# Unravelling the joint genetic architecture between psychiatic and insulin-related traits in the general population

**DOI:** 10.1101/2024.10.04.24314905

**Authors:** Barbara Sakic, Izel Erdogan, Giuseppe Fanelli, Martina Arenella, Nina Roth Mota, Janita Bralten

## Abstract

Attention-deficit/hyperactivity disorder (ADHD), autism spectrum disorder (ASD), and obsessive-compulsive disorder (OCD) are heritable disorders that frequently co-occur with insulin resistance (IR)-related conditions. Traditional genetic case-control comparisons are challenged by the extent of heterogeneity and comorbidity within and across these conditions. In this study we step away from univariate analyses to let biology guide us to the potential genetic links between insulin and psychiatry-related traits.

We used large-scale population-based genetic studies (N= 17,666-697,734) and applied genomic structural equation modeling to identify the factor structure best representing the joint genetic architecture of symptom scores of ADHD, ASD, and OCD, and five IR-related traits: body mass index (BMI), fasting plasma glucose (FPG), fasting plasma insulin, glycated haemoglobin (HbA1c), and homeostatic model assessment for IR. Subsequently we performed multivariate genome-wide association analyses on the psychiatry-IR related factors to explore genetic associations to unravel its biological basis. Factor analyses indicated that a three-factor model fitted the data best (x2(df=9)=18.79, AIC=56.8, CFI=0.99, SRMR=0.068). One factor included ADHD traits and three IR-related traits (BMI, FPG, HbA1c), while another encompassed OCD traits and HbA1c. The last factor included solely IR-related traits. Gene-wide analyses revealed 57 genes significantly associated with the ADHD-IR factor (p< 2.961e-06) and one gene, *MTNR1B* (p=3.44e-07), with the OCD/OCS-IR factor. Gene-set analyses found associations with neurodevelopmental pathways.

Our findings suggest a shared genetic liability between psychiatric symptoms and IR-related traits in the general population, offering new perspectives on the molecular genetics underlying the overlap between psychiatric and IR-related somatic conditions as well as biologically informed clustering within psychiatry.

## Introduction

Attention-deficit/hyperactivity disorder (ADHD), autism spectrum disorder (ASD), and obsessive-compulsive disorder (OCD) are common neurodevelopmental disorders that frequently co-occur[1,2]. These complex psychiatric disorders are known to be heritable and have complex multifactorial etiologies [3,4,5]. Univariate genome-wide association studies (GWAS) comparing individuals with ADHD, ASD or OCD to those without these disorders have identified the first disorder-associated common genetic variants [6,7,8]. However, enormous samples sizes were needed to find single variants with small effect sizes and underlying biological mechanisms still remain largely unknown. One challenge in gene finding is the observed heterogeneity of clinical manifestations in psychiatric disorders, where psychiatric patients with the same diagnostic label can differ substantially from each other. Currently, clinical diagnoses for ADHD, ASD, and OCD are dependent on arbitrary threshold of symptoms only, without a biological basis [9,10]. Due to the lack of biological understanding, ambiguous diagnoses and large amounts of co-morbidity exist in individuals with psychiatric disorders, which extends beyond psychiatric phenotypes and also includes insulin resistance (IR)-related somatic conditions, like type 2 diabetes mellitus (T2D) and obesity [11,12,13].

The link between ADHD, ASD, and OCD and IR-related somatic conditions is supported by epidemiological data. It has been found that having T2D or other metabolic conditions during pregnancy increases the risk of the offspring developing ASD or ADHD [14,15,16]. In addition, large-scale registry data convincingly showed that obesity is a relevant risk factor not only for developing T2D and metabolic syndrome, but also for receiving mental health diagnoses [17] and that bidirectional associations exist between T2D and psychiatric disorders, including ADHD, ASD, and OCD [18]. The observed multimorbidity between IR-related conditions and psychiatric disorders complicates clinical trajectories [19] and have been linked to more severe clinical outcomes [20,21] indicating their potential clinical relevance.

One connection between psychiatric conditions and IR-related somatic conditions might lie in dysregulated insulin. While the role of insulin in glycemic control in our body is well known, it is becoming more clear that insulin also has an important function in the brain [22]. In the brain, insulin activates insulin receptors expressed on the surface of neuronal and glial cells, initiating a cascade of intracellular transduction processes [23]. By mediating cellular metabolism, insulin contributes to the development and homeostasis of the central nervous system, influencing neurogenesis, neuronal differentiation and promoting neurite growth [24]. Additionally, insulin has protective properties in the brain preventing damage from apoptosis and oxidative stress [24]. Previous studies show that IR-related conditions and traits associated with cognitive performance including IR-related associations with poorer verbal and numerical reasoning ability and slower processing speed [25] and global cognitive function [26]. While metabolic disturbances in psychiatry have been perceived as possible consequences of unhealthy lifestyles, sedentary habits, or the chronic use of psychotropic medication [27], the observation that glycaemic and metabolic imbalances have been found in drug-naïve acute psychiatric patients already at disease onset suggest the potential involvement of common biological mechanisms [28]. Until now, our understanding of these underlying mechanisms linked to the multimorbidity of psychiatric disorders and IR-related conditions is limited.

Since IR-related somatic diseases are complex heritable diseases as well [29], like ADHD, ASD, and OCD, one way to learn more about potential shared etiologies could lie in genetic analyses. It was already noted in GWAS investigating OCD that genes regulating insulin signaling were enriched in the top results [30] and that insulin-related traits, including levels of fasting insulin and two hour glucose measures, showed significant genetic correlations with OCD [31]. Genetic sharing between IR-related conditions [32] and psychiatric disorders [33] have also been well reported. A recent study investigated the genetic cross-links between nine neuropsychiatric disorders and three IR-related somatic diseases using publicly available GWAS data [34]. They were able to show two distinct clusters, in which the genetics of IR-related conditions may exert divergent pleiotropic effects: one including OCD as well as anorexia nervosa and schizophrenia, which showed negative genetic overlap with somatic IR-related conditions, and the other one including ADHD and major depressive disorder showing positive genetic overlap with IR-related conditions [34]. In addition, a recent family-based study was able to show that relatives of individuals with a psychiatric disorder had an increased risk for T2D and, within a subpart of their study, they were able to show that genetic risk for T2D was associated with an increased risk for several psychiatric disorders, including ASD and ADHD [35]. These studies highlight the evidence for multiple genetic links between IR-related somatic diseases and ADHD, ASD, and OCD.

Another challenge in gene finding in psychiatry, apart from the phenotypic heterogeneity, lies in genetic heterogeneity. Genetic evidence shows that psychiatric disorders share genetic risk factors [36], and cross-disorder studies have identified pleiotropic genetic risk factors involved in several psychiatric disorders [37]. These results indicate that the clinical diagnoses do not follow the biology well, and first studies now indicate that substructures might exist in the genetic contributions to psychiatric categories [38,39]. With examples like T2D being prevalent in 5-22% of individuals with psychiatric disorders [40] and the other way around, with one-third to almost two-thirds of somatic patients having mental disorders [41], IR could be an interesting starting point to explore the underlying biology of heterogeneous psychiatric phenotypes. Moreover, the extensive genetic links between IR-related conditions and psychiatric conditions also support this interest. Furthermore, somatic IR-related traits can be objectively measured, potentially allowing for the stratification of individuals based on objectively measurable features, something we currently still lack in clinical psychiatry.

With classical genetic case-control comparisons being challenged by the extend of heterogeneity and comorbidity within and across psychiatric conditions, researchers have explored alternative ways to study psychiatric genetics. One of these approaches is to explore continuous phenotypes [42,43]. Many symptoms/characteristics of psychiatric disorders are also observed in healthy individuals. Such disorder-like traits tend to show a nearly normal distribution in the general population, with psychiatric cases clustering at the extreme end [44]. It was shown that this trait-disorder continuum also holds at the genetic level, demonstrated by genetic overlap between obsessive-compulsive traits (’guilty taboo thoughts’) and OCD [45, 46], autistic-like traits (including social withdrawal) and ASD [47,48] and attention-deficit/hyperactivity symptoms and ADHD [49]. Additionally, some traits span multiple diagnoses, indicating trans-diagnostic characteristics that align with the genetic correlations found in cross-disorder studies [33]. Investigating these continuous traits can help elucidate the observed heterogeneity in clinical phenotypes.

While bi-variate genetic analyses have been instrumental for showing shared genetic etiologies between psychiatric traits and IR-related traits, new statistical tools make it possible to identify genetic relationships associated with multiple traits. Genomic Structural Equation Modelling (Genomic SEM) is a multivariate method that can analyze the joint genetic architecture of complex traits [50]. First results indicate patterns of substructures in the genetic correlations between major psychiatric disorders and show that multivariate genetic analyses are able to find additional genetic risk variants that were not found in univariate analyses [51,52]. This study aims to explore the existence of latent genetic factors based on the genetics of ADHD, ASD, and OCD symptomatology, as well as IR-related continuous measures observed in population-based genetic studies to examine its joint genetic architecture. We leveraged trait-based GWAS focusing on the continuum of symptoms related to ADHD, ASD, and OCD rather than the differences between cases and controls. We applied multivariate genomic SEM to parse heterogeneity and genetic overlap between these psychiatric phenotypes and IR-related traits. Subsequently, we performed multivariate GWASs on the modeled genetic factors to uncover genetic and biological underpinnings of the latent genetic factors representing shared genetics between psychiatric and IR-related traits.

## Materials and Methods

### Data source

We utilized publicly available summary statistics [53] from large-scale population-based GWASs that included symptoms related to ADHD, ASD, OCD or IR-related measures (N=17,666 - 697,734). The inclusion criteria for GWASs consisted of a minimum sample size of 10 000 and having significant SNP-based heritability (p<0.05; Z>2). To ensure the inclusion of the largest possible GWAS datasets available, we utilized the online database GWAS Catalog [53] for the final selection. An overview of GWAS datasets used is provided in Table 1.

**Table 1.**
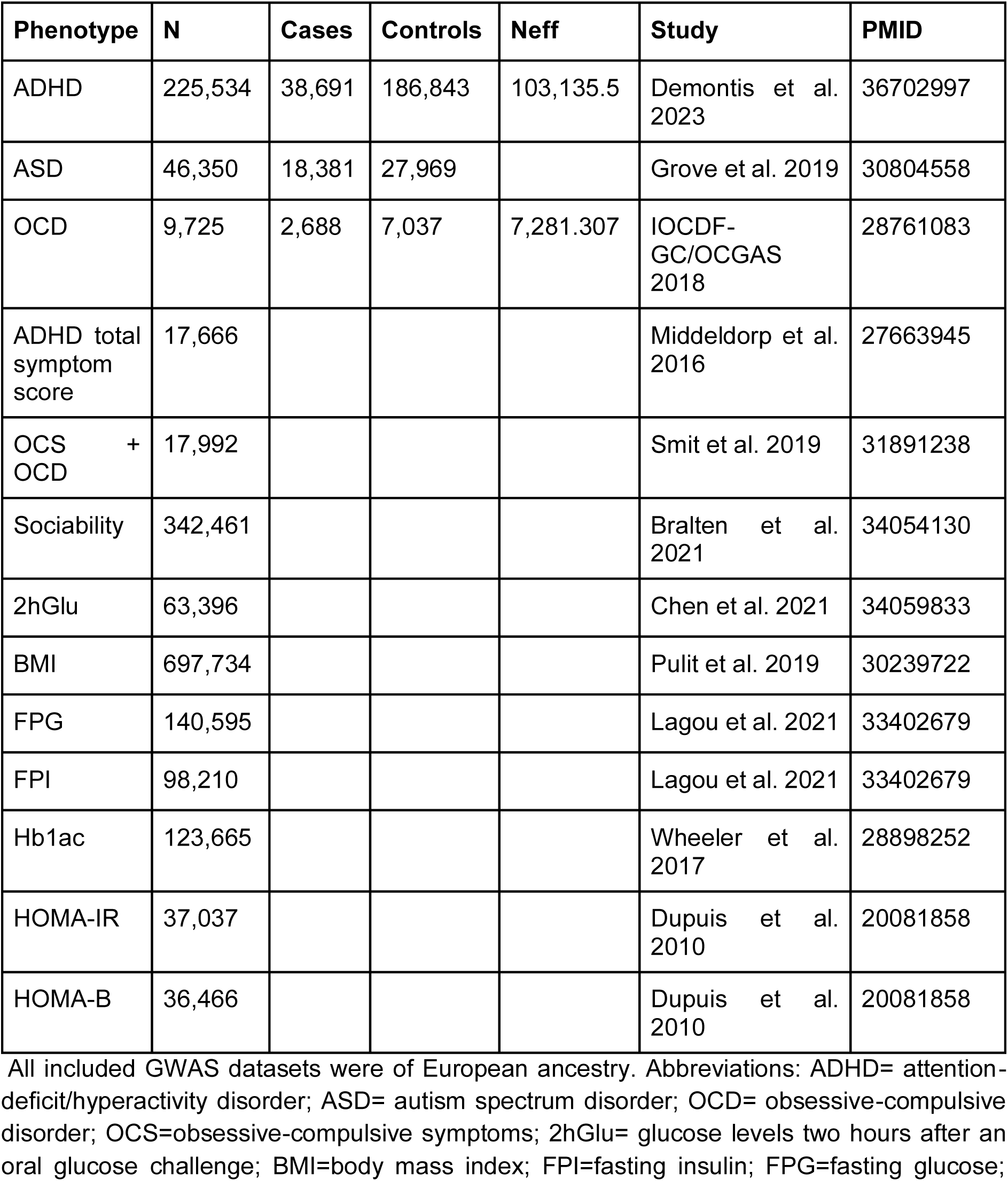
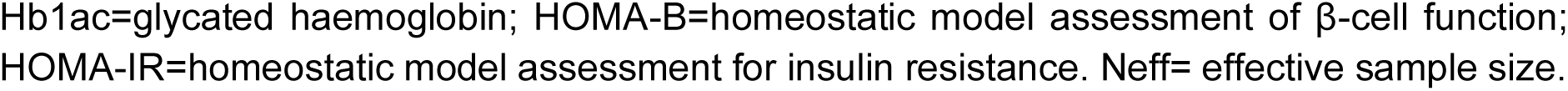
Overview of included GWAS datasets.

| Phenotype | N | Cases | Controls | Neff | Study | PMID |
| --- | --- | --- | --- | --- | --- | --- |
| ADHD | 225,534 | 38,691 | 186,843 | 103,135.5 | Demontis et al. 2023 | 36702997 |
| ASD | 46,350 | 18,381 | 27,969 |  | Grove et al. 2019 | 30804558 |
| OCD | 9,725 | 2,688 | 7,037 | 7,281.307 | IOCDF-GC/OCGAS 2018 | 28761083 |
| ADHD total symptom score | 17,666 |  |  |  | Middeldorp et al. 2016 | 27663945 |
| OCS + OCD | 17,992 |  |  |  | Smit et al. 2019 | 31891238 |
| Sociability | 342,461 |  |  |  | Bralten et al. 2021 | 34054130 |
| 2hGlu | 63,396 |  |  |  | Chen et al. 2021 | 34059833 |
| BMI | 697,734 |  |  |  | Pulit et al. 2019 | 30239722 |
| FPG | 140,595 |  |  |  | Lagou et al. 2021 | 33402679 |
| FPI | 98,210 |  |  |  | Lagou et al. 2021 | 33402679 |
| Hb1ac | 123,665 |  |  |  | Wheeler et al. 2017 | 28898252 |
| HOMA-IR | 37,037 |  |  |  | Dupuis et al. 2010 | 20081858 |
| HOMA-B | 36,466 |  |  |  | Dupuis et al. 2010 | 20081858 |

All included GWAS datasets were of European ancestry. Abbreviations: ADHD= attention-deficit/hyperactivity disorder; ASD= autism spectrum disorder; OCD= obsessive-compulsive disorder; OCS=obsessive-compulsive symptoms; 2hGlu= glucose levels two hours after an oral glucose challenge; BMI=body mass index; FPI=fasting insulin; FPG=fasting glucose;
Hb1ac=glycated haemoglobin; HOMA-B=homeostatic model assessment of β-cell function; HOMA-IR=homeostatic model assessment for insulin resistance. Neff= effective sample size.

### GWASs of population-based psychiatric traits

To include a population-based trait for ADHD, we considered the GWAS meta-analysis of a total score of attention and hyperactivity symptoms in children [49]. This meta-analysis includes 17,666 children from nine population-based cohorts of the EArly Genetics and Life course Epidemiology (EAGLE) consortium. Data were based on scaled questionnaires, including the Attention Problems scale of the Child Behavior Checklist (CBCL) and the Hyperactivity scale of the Strengths and Difficulties Questionnaire (SDQ), consisting of parent- and teacher-rated scales [49].

For OCD traits, previous GWASs of obsessive-compulsive symptoms did not reach the sample size requirements for Genomic SEM. Therefore, we used a GWAS that meta-analyzed obsessive-compulsive symptoms (OCS) in combination with OCD case-control status [54]. The OCS score in this meta-analysis used subscales that measured obsessions (rumination and impulsions) and compulsions (checking, washing, and ordering/precision), based on the Padua Inventory scale [54]. The continuous OCS scores were measured in a population with an age range between 18 and 80 years, including 8,267 individuals based on self-reported items. The OCD case-control subsample included 2,688 individuals with OCD and 7,037 controls from the Psychiatric Genomics Consortium OCD GWAS [8].

For ASD-related traits, we used a GWAS on sociability [48] since social difficulties are at the core of the autistic phenotype and there is evidence of genetic overlap between sociability and ASD genetics [48]. The sociability GWAS was performed on an aggregated score based on four questions related to social behavior in a total of 342,461 adults (mean age 56.61). Further information regarding the sociability measure items and genotyping has been reported [48]. In our analysis, the direction of beta association values of SNPs from the sociability GWAS summary statistics was reversed to indicate reduced sociability, reflecting social withdrawal.

### GWASs of population-based IR-related traits

GWAS data on IR-related traits were preselected based on the aforementioned inclusion criteria, as well as the existence of significant genetic correlations with either ADHD, ASD, or OCD, aligning with Genomic SEM requirements. IR-related traits considered were: glucose levels two hours after an oral glucose challenge (2hGlu), body mass index (BMI), fasting glucose (FPG), fasting insulin (FPI), glycated haemoglobin (HbA1c), homeostatic model assessment for insulin resistance (HOMA-IR), and homeostatic model assessment of β-cell function (HOMA-B) [55,56,57,58,59]. We performed genetic correlation analyses with linkage disequilibrium score (LDSC) regression between the IR-related traits with psychiatric disorders (ADHD, ASD, OCD) as described in Supplementary Materials in Supplementary Figure S1 and Table S1.

### Genomic Structural Equation Modeling

Genomic SEM was applied to investigate latent genetic factors underlying the three population-based symptom GWASs (attention-deficit/hyperactivity symptom scores, OCD+OCS symptoms, social withdrawal) and the five IR-related trait GWASs (BMI, FPG, FPI, HbA1c, HOMA-IR) that were significantly genetically correlated to at least one among ADHD, ASD, and OCD. Genomic SEM is a data-driven, multivariate genetic analysis approach used to identify latent factors underlying different variables [50]. The analysis involves exploratory and confirmatory factor analyses (EFA and CFA, respectively) to determine the model based on loading thresholds above 0.25 and thereafter examine model fit. Factor analyses were performed separately for the odd and even chromosomes to avoid overfitting. In the first step of Genomic SEM, the genetic covariance matrix and sampling covariance matrix were estimated using precomputed linkage disequilibrium (LD) scores obtained from the 1000 Genomes Project [60]. During the second step, the model fit was estimated using standardized root mean square residual (SRMR), model x2, Akaike Information Criterion (AIC), and the Comparative Fit Index (CFI). The model fit statistic CFI tests to what degree the proposed model better fits compared to a model in which all traits are heritable but not correlated genetically (>.95 is considered good fit), and AIC is a relative fit statistic to compare models, with lower values being a better fit [50]. The analysis followed procedures described on the Genomic SEM GitHub, using default parameters (see https://github.com/GenomicSEM/GenomicSEM).

### Multivariate GWAS

We performed multivariate GWASs within the Genomic SEM framework on the factors that loaded both psychiatric and IR-related traits to estimate individual SNP effects on the latent factors. Multivariate GWAS was run on SNPs filtered on default SNP filtering on minor allele frequency (MAF) (MAF>0.01) and quality info score (INFO>0.6) when such information was available [51]. In addition, we removed any variants that presented additional warnings during the analysis, such as NA values. SNPs were considered significant if they reached the genome-wide significance threshold of p <5e-08. A SNP-level heterogeneity test (QSNP) was then performed to test the null hypothesis that a SNP acts on the latent factor instead of acting on individual or subset of traits/observed variables. Therefore, we calculated the heterogeneity (Q) index [50, 51] by using independent pathway models in which the effects of the SNPs on the traits and the residual variances are freely estimated. The common and independent pathway models are then used to estimate a x2 distributed QSNP test statistic with degrees of freedom (df) equal to k-1, where k reflects the number of included phenotypes. We then removed significant QSNPs (p< 5e -08) for subsequent analyses as these SNPs do not represent direct effects on latent factors.

The effective sample size (Neff) for each psychiatric-IR factor was calculated [50, 61] and in this calculation the summary statistics are restricted to MAF limits of 10% and 40% to produce stable estimates, as described on the Genomic SEM Github (https://github.com/GenomicSEM/GenomicSEM/wiki/5.-User-Specified-Models-with-SNP-Effects).

### FUMA

The online tool for functional mapping and annotation of GWASs (FUMA v1.5.4) [62] was utilized to interpret the multivariate GWAS outcomes of each psychiatric-IR related latent factor. The SNP2GENE module within FUMA was used to perform gene mapping (i.e., positional mapping, eQTL mapping), as well as gene-based, gene-property, and gene set analyses in Multi-marker Analysis of GenoMic Annotation (MAGMA). The gene-wide genome-wide significance threshold was set at p=0.05/16 888=2.961e-06 accounting for the 16,888 tested protein coding genes. The gene set analysis was performed on 5,917 Gene Ontology (GO) terms and 4,761 curated gene sets obtained from MsigDB v6.2. Gene set analysis results were considered significant at p<0.05/10 678=4.68e-06. For the gene-property analyses we selected 30 GTEx/v8, 53 GTEx/v8 and BrainSpan gene expression datasets to examine gene expression in brain-related regions in adults and during brain development. As input for the sample size parameter (N) in FUMA we used the Neff due to inputting multivariate GWAS outcomes in FUMA analysis.

## Results

### Identification of psychiatric-IR latent genetic factors

Based on our input data of eight variables: three psychiatric traits (ADHD total symptom score, OCS and OCD case-control score, and social withdrawal) and five IR-related continuous traits (BMI, FPI, FPG, Hb1ac, HOMA-IR) the EFA analysis within Genomic SEM on odd chromosomes provided four potential models. The subsequent CFA analysis on these models on even chromosomes confirmed the best model fit for a three-factor model (x2(df=9)=22.87, AIC=60.8, CFI=0.97, SRMR=0.093), see Figure 1. This model also fits well for all chromosomes (x2(df=9)=18.79, AIC=56.8, CFI=0.99, SRMR=0.068).

**Figure 1.**
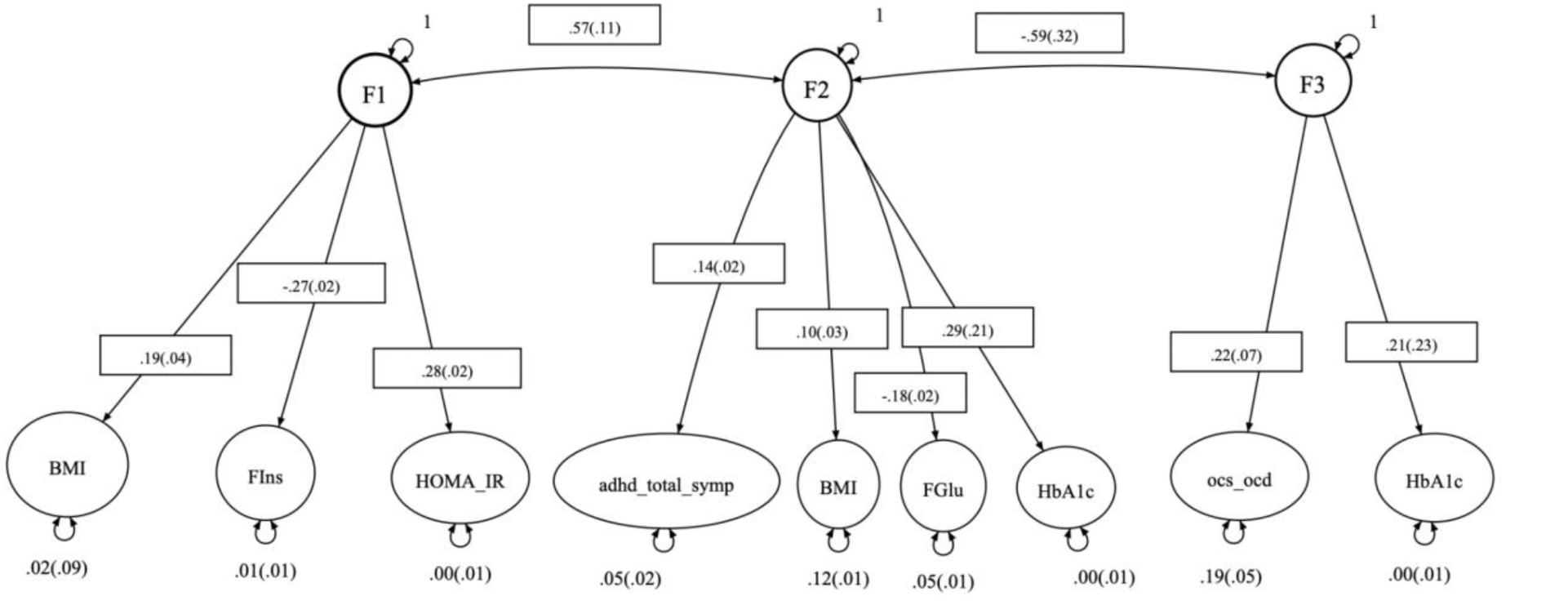
Three-factor model path diagram. The structural equation model path diagram of the identified three-model as the best model fit to the data based on the exploratory factor analysis in Genomic SEM. Abbreviations: adhd_total_symp= ADHD total symptom score; ocs_ocd= obsessive-compulsive and OCD case-control score; BMI=body mass index; FPI=fasting insulin; FPG=fasting glucose; Hb1ac=glycated haemoglobin; HOMA-IR=homeostatic model assessment for insulin resistance. Created with https://semdiag.psychstat.org/.

The first factor was loaded solely by IR-related traits (BMI, FPI, and HOMA-IR). The second factor included ADHD total symptom scores together with BMI, FPG, and HbA1c and was named ADHD-IR factor. The third factor included OCD+OCS symptoms together with HbA1c and was named OCD/OCS-IR factor. Social withdrawal did not load in any of the three factors.

### SNP associations

Multivariate GWAS were performed on the ADHD-IR and the OCD/OCS-IR factors. Multivariate genome-wide associations on the ADHD-IR factor identified two genome-wide significant SNPs, rs780093 (p=1.56e-08) and rs551754 (p=1.63e-08) (See Supplementary Figure S2). Heterogeneity tests indicated a total of 1,304 heterogeneous QSNPs, and both genome-wide significant associations were QSNPs and therefore removed for subsequent analyses (See Supplementary Figure S3). The analysis identified no genome-wide significant associations for the OCD/OCS-IR latent factor (See Supplementary Figures S4 and S5) and 813 heterogeneous QSNPs, which were removed for subsequent analyses. The most significant SNPs for the ADHD-IR factor and OCD/OCS-IR factors provided by FUMA can be found in Supplementary Tables S2 and S4. Additional quality control statistics (quantile-quantile plots) are available in Supplementary Figures S6 and S7.

### Gene-level associations

Using the FUMA SNP2GENES module, SNPs from the multivariate GWAS outputs of the ADHD-IR and OCD/OCS-IR latent factors were annotated to 16,888 protein coding genes. The built-in MAGMA gene-based analysis identified 57 genes reaching genome-wide significance for the association with the ADHD-IR factor (Figure 2 and Supplementary Table S3), and one gene, *MTNR1B* gene (p=3.4364e-07), for the OCD/OCS-IR factor (Figure 3).

**Figure 2.**
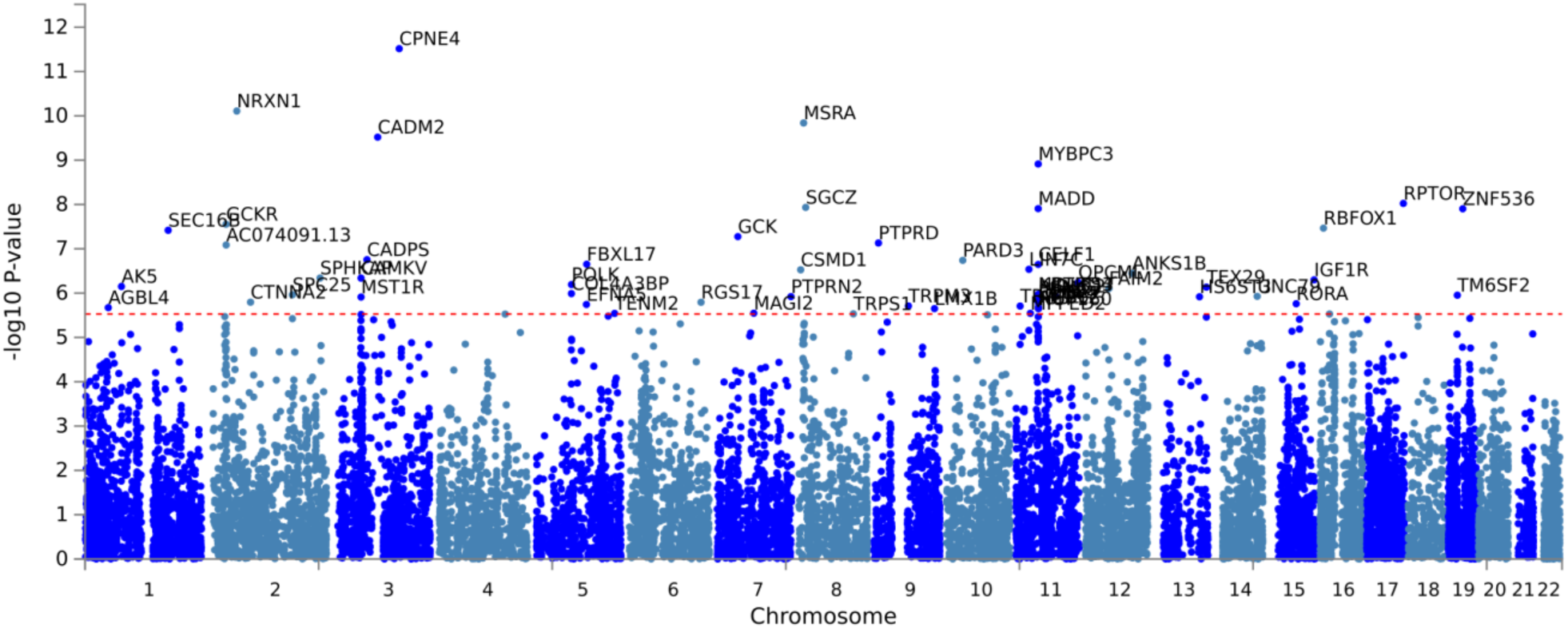
Manhattan plot of gene-based analysis of ADHD-IR latent factor. Gene-based Manhattan plot based on the input GWAS summary statistics of the multivariate GWAS of the ADHD-IR factor. The input SNPs of the GWAS summary statistics were mapped to a total of 16888 protein coding genes. The x-axis displays the positional location on the different chromosomes, and the y-axis displays the -log p-value of the gene-wide associations. The genome wide significance threshold (red dashed line in the plot) was set at p=0.05/16888=2.961e-6 accounting for the number of genes tested. The plot shows the 57 significant top genes.

**Figure 3.**
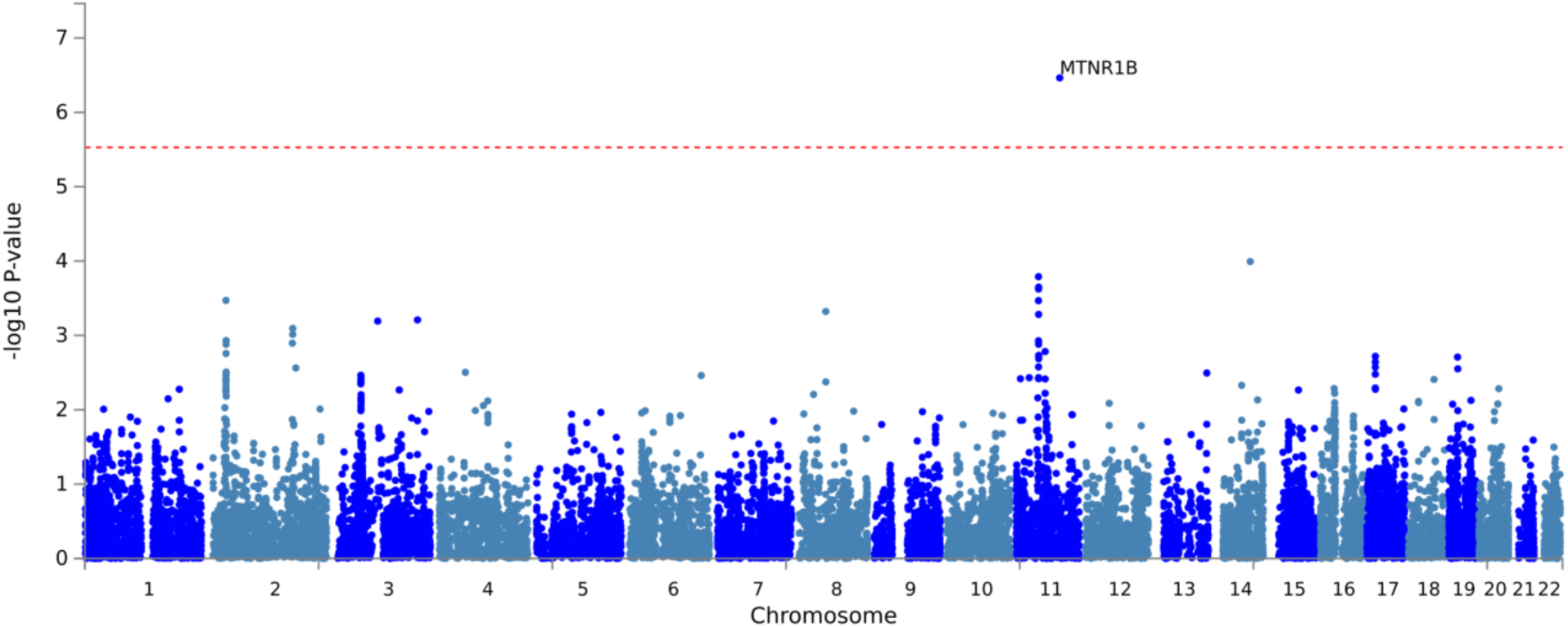
Manhattan plot of gene-based analysis of OCD/OCS-IR latent factor. Gene-based Manhattan plot based on the input GWAS summary statistics of the multivariate GWAS of the OCD/OCS-IR latent factor. The input SNPs of the GWAS summary statistics were mapped to a total of 16884 protein coding genes. The x-axis displays the positional location on the different chromosomes, and the y-axis displays the -log p-value of the gene-wide associations. The genome wide significance threshold (red dashed line in the plot) was set at p=0.05/16884=2.961e-6. accounting for the number of genes tested.

### Gene set enrichment and tissue specificity

Subsequently, MAGMA gene-set analysis allowed to explore factor-associated gene sets. This analytical step yielded significant results for 12 gene sets associated with the ADHD-IR factor and five gene ontology terms associated with the OCD/OCS-IR factor (Table 2 and 3, respectively), including neuron differentiation, neuron development and neurogenesis.

**Table 2.**
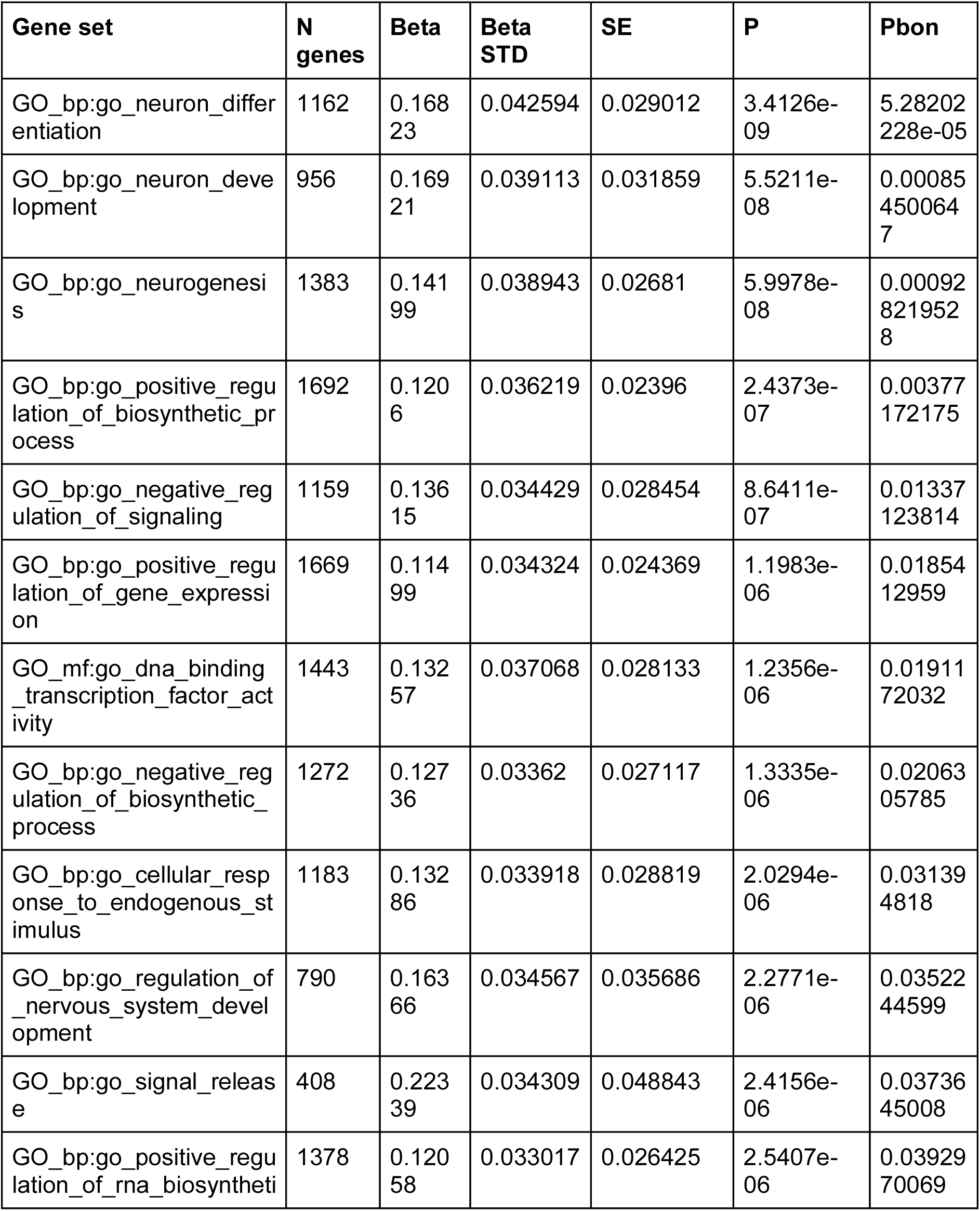
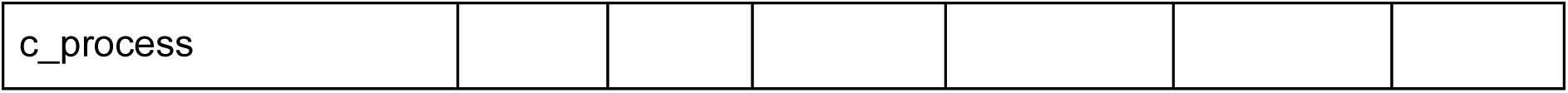
Results of the MAGMA gene set based analysis of ADHD-IR latent factor. The table shows the top significant identified gene sets based on Bonferroni corrected p-values accounting for 5917 Gene Ontology (GO) terms and 4761 curated gene sets. N genes= number of genes; Beta=beta value; Beta STD=beta value standard deviation; SE=standard error; P=p-value ; Pbon=p-value after Bonferroni correction.

**Table 3.**
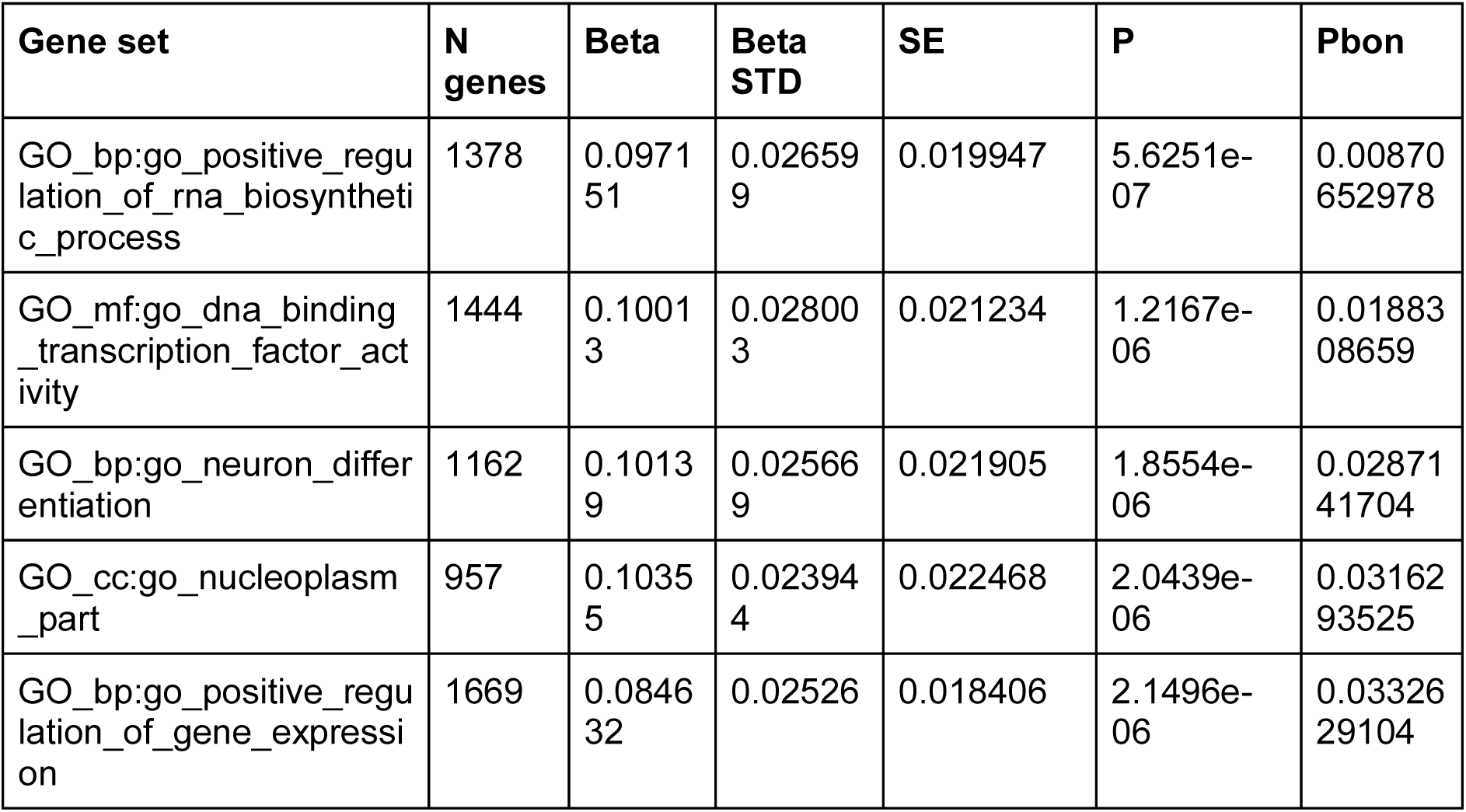
Results of the MAGMA gene set based analysis of OCD/OCS-IR latent factor. The table shows the top significant identified gene sets based on Bonferroni corrected p-values accounting for 5917 Gene Ontology (GO) terms and 4761 curated gene sets. N genes= number of genes; Beta=beta value; Beta STD=beta value standard deviation; SE=standard error; P=p-value ; Pbon=p-value after Bonferroni correction.

Gene-property analyses indicated significant enrichments in the brain and pituitary for genes related to the ADHD-IR factor across 30 general tissue types. Moreover, it indicated significant enrichments in all brain regions, including cerebellum, except the substantia nigra for genes related to the ADHD-IR factor across 53 tissue types. Additionally, significant enrichments were found between early and late mid-prenatal developmental stages of the brain for genes related to the ADHD-IR factor (Supplementary Figure S8). For the OCD/OCS-IR factor, we observed significant enrichments in the brain, pituitary and uterus across 30 general tissue types and seven out of the 13 brain regions across 53 specific tissue types. In addition, significant enrichments were observed from early prenatal to late mid-prenatal developmental stages of the brain for genes related to the OCD/OCS-IR factor (Supplementary Figure S9).

## Discussion

Psychiatric phenotypes and IR-related conditions co-occur and are genetically correlated, yet the molecular genetic underpinnings of this overlap remain poorly understood. This study employed multivariate genomic analyses of population-based traits to demonstrate the existence of genetic factors that include ADHD symptoms and IR-related measures, and obsessive-compulsive symptoms and cases, and IR-related measures. Follow-up bioinformatic analyses suggest the potential involvement of IR-related genes as well as genes involved in neuron differentiation, neuron development, and neurogenesis. Identifying these psychiatric-IR genetic factors may be a step towards biologically informed subgroups within psychiatry in the future. Our work shows the existence of a latent genetic factor loaded by ADHD total symptom scores and three IR-related traits (BMI, FPG, and HbA1c). These IR-related traits have previously shown positive genetic correlations with ADHD [34] and results are in line with phenotypic observations of elevated HbA1c levels in children with ADHD compared to children without ADHD [63]. Poor glycaemic control within diabetic patients has been linked to increased risk of ADHD [64], suggesting the potential of a distinct biological profile for a subgroup of individuals with ADHD. In addition, our results are in line with a previous study describing individuals diagnosed with T2D being susceptible to presenting ADHD-related symptoms [65]. Previous work also indicates a link between insulin and dopamine signaling in the midbrain making a connection to altered reward behavior [66]. Altered reward sensitivity is known in ADHD [67] and dopamine and ADHD have been linked through different routes, including genetics [68], brain imaging [69], as well as the pharmacodynamics of symptom reducing medications [70].

We also identified a latent genetic factor loading OCD/OCS and HbA1c. An IR-related OCD/OCS factor is in line with studies linking insulin to OCD. Among these, Hou and colleagues found that individuals diagnosed with OCD present increased glucose metabolism in the orbitofrontal cortex compared to healthy controls [71], aligning with rodent models of T2D that presented compulsivity-related behavior, and increased glucose levels in the dorsomedial striatum [72]. Also genetic work has linked insulin genes to both OCD [30] and OCS [31]. For HbA1c no genetic sharing with OCD measures was observed previously [31]. Nonetheless, HbA1c levels have been shown to be positively correlated to OCD symptomatology [73,74]. Prior work did report genetic sharing between OCD and FPI levels and the 2hGlu measure [31], while negative genetic correlations of BMI and obesity have been described with OCD diagnosis [34]. Strom and colleagues defined OCD subgroups based on comorbid diagnoses and genetic relations between psychiatric disorders with somatic and mental measures. Their study showed that polygenic risk score for BMI was more negatively associated with the OCD group without comorbidities compared to OCD subgroups having a comorbid condition, such as ADHD [75]. This observation may highlight the specific negative genetic relationship between OCD and the IR-related trait BMI. The disparate genetic relationships of OCD versus ADHD to different traits, specifically IR-related traits, denotes potential different underlying mechanisms. Again, this is in line with the defined clusters related to IR-related multimorbidity observed, placing ADHD and OCD in different clusters [34].

The results of our multivariate genetic analyses highlighted 57 genes significantly associated with the ADHD-IR factor, including *CPNE4*, encoding copine 4. Copines are a family of calcium-dependent membrane binding proteins enriched in neurons [76] and involved in insulin secretion and glucose uptake regulation [77]. Another top gene was *CADM2*, encoding cell adhesion molecule 2. *CADM2* has been linked to core ADHD features like impulsivity and risk-taking behavior [78]. Conditional genetic analyses showed that variants within *CADM2* influence both psychiatric and metabolic traits, including BMI, and are expressed in adult brain and adipose tissue [79], suggesting common biological mechanisms. Interventions on expression of *CADM2* in animal models resulted in changes in adiposity, systemic glucose levels and insulin sensitivity [80], linking this gene to glycemic regulation. This insulin-psychiatric link is also apparent in other top genes, including *NRXN1*, implicated in insulin vesicle granule docking [81] and neurodevelopmental disorders, including ASD and depression [82,83,84]. Hughes and colleagues investigated an ASD mouse model with reduced Nrxn1a expression, that showed increased glucose metabolism in the dorsal raphe nucleus, and decreased insulin receptor signaling in the prefrontal cortex, a top identified brain region in our gene property analysis for the ADHD-IR factor associated genes [84]. We also find *RPTOR* as a top associated gene to the ADHD-IR genetic factor. This gene encodes the regulatory-associated protein of mTOR, or raptor, implicated in the mTOR pathway and regulating cell growth and survival [85]. The genetic variants within the mTOR pathway has been described as a link between brain volume and ASD in earlier analyses [86]. This regulatory protein is sensitive to insulin levels, and also regulates glucose metabolism and synaptic plasticity via the mTOR cascade [87]. Moreover, upregulation of mTOR-related genes has not only been observed in ASD, but also in ADHD [87].

Next to single genes we also observed top significant associated gene sets with the ADHD-IR factor that included neuron differentiation, neuron development, and neurogenesis in accordance with ADHD being a neurodevelopmental disorder [88]. Based on accumulating evidence for the actions of insulin in the brain [24,89], we could speculate that the role of insulin in neurodevelopment could be important in relation to our identified factor. In tissue expression analyses of the genes associated to the ADHD-IR latent factor, we observed significant enrichments in all tested brain regions except the pituitary and the substantia nigra. The positive association between gene expression in the cerebellum and genetic associations of the ADHD-IR factor link to the finding of a previous study reporting an association between smaller volume of the vermis of the cerebellum and the amount of ADHD symptoms [90]. ADHD symptoms scores on an attention scale also associated to smaller surface areas, including frontal gyrus and total surface area, in a pediatric population cohort [91]. Our findings linking gene expression in the frontal cortex BA9 region and brain cortex are in line with previous work suggesting aberrated maturation of frontal lobes may result in ADHD symptoms in a subgroup of children with ADHD [92].

Children with ADHD have increased waist circumference and BMI compared to children without ADHD, and these measures are linked to the severity of the condition [93]. Children with ADHD also show unhealthy dietary behavior, consuming less vitamins and simple sugars in contrast to undiagnosed children [93]. Such observations, along with our results, may direct to new avenues to identify subgroups of individuals with attention-deficit/hyperactivity symptoms who may benefit from receiving personalized lifestyle-based interventions.

We identified *MTNR1B* as a genome-wide significant gene associated to the OCD/OCS-IR factor. This gene encodes the melatonin receptor 1B, a high affinity membrane receptor for the melatonin hormone [94]. *MTNR1B* is implicated in glucose homeostasis due to melatonin functioning, as elevated melatonin levels may impair glucose tolerance and modulate fasting glucose levels [95], linking this gene to glycemic regulation. Altered levels of melatonin have been associated to the risk for type 2 diabetes as well as disturbed circadian rhythm [94,96]. Lane and colleagues investigated circadian rhythm based on sleep measures via self-reports and physiological measurements with polysomnography. Here, they reported that circadian disruption may mediate risk for type 2 diabetes via *MTNR1B* [94]. Sleep disturbances are also described in OCD, linking the severity of obsessive-compulsive symptom severity to sleep quality [97]. Finding neuron differentiation in our gene set analysis links neuronal development to the genetic architecture of the OCD/OCS-IR factor, in line with the known neurodevelopmental trajectory of OCD [8].

The OCD/OCS-IR factor related genes were associated with gene expression across a total of 7 brain regions, including the brain cerebellar hemisphere and brain cerebellum. Links between OCD and the brain have been reported before. Zhang and colleagues reported an association between functional connectivity between posterior cerebellum and right striatum, and OCD symptom severity in individuals diagnosed with OCD [98]. We reported positive associations between the mid-prenatal and prenatal developmental stages of the brain and the genetic associations of the OCD-IR-related factor, linking the OCD/OCS-IR genetic factor to fetal brain development. A similar conclusion was reached regarding OCD and fetal origin based on BrainSpan transcriptome profiles in a supervised learning approach study [99], also supporting the link of prenatal developmental trajectory to our genetic factor.

We hypothesized a role of insulin signaling in ASD but did not identify a genetic factor including our ASD-related trait social withdrawal and the IR-related traits. While previous work also did find neither global nor local genetic correlations between IR-related measures and ASD [34,100], stratified analyses did indicate a significant covariance for genes within the insulin signaling pathway between ASD and metabolic syndrome [34]. It must be noted that an ASD diagnosis consists of more than social withdrawal-related symptoms, however in the current analyses we restricted to social withdrawal because of its adequate sample size as there was no total autism spectrum-like symptom GWAS available with a sufficient sample size. Therefore, we cannot conclude the absence of the existence of a potential ASD-related IR genetic factor, but we can only report not finding one with social withdrawal.

This study has several strengths and limitations. By moving beyond a traditional case-control approach to the study of population-based psychiatric and IR-related traits, we yielded novel insights into the biological basis of symptom dimensions in psychiatry, while gaining more statistical power through the use of large input GWAS datasets in our analyses. Genetically-based factors, as demonstrated by the identified ADHD-IR and OCD/OCS-IR genetic clusters, can delineate biologically informed grouping. Moreover, our study highlights the utility of integrating psychiatric and somatic traits to better understand the complex interplay between these factors. However, our study also has some limitations. The reliance on publicly available population-based GWAS data introduces dependence on current data availability. The limited sample size of some studies may impede the discovery of genetic variants due to insufficient statistical power. Additionally, the use of European cohorts hinders the generalizability of our findings to other ethnicities. Future research should examine if our findings can be validated in clinical settings and explore if the biological clustering can be observed in diverse populations.

In conclusion, this study identified latent genetic factors characterized by shared liabilities underlying ADHD symptoms with IR-related measures and obsessive-compulsive symptoms and OCD cases with IR-related measure. The ADHD-IR factor included ADHD total symptom scores with BMI, FPG, and HbA1c, while the OCD/OCS-IR factor comprised OCS and OCD traits with HbA1c. These factors could potentially aid in pointing towards subgrouping within psychiatric disorders like ADHD and OCD, characterized by specific IR-related profiles. To our knowledge, this is the first study reporting on the multivariate genetic clustering of dimensional psychiatric symptoms with somatic IR-related traits in population-based datasets. Our work highlights the shared genetic architecture of psychiatric and IR-related traits, and provides a starting point for the potential of clustering individuals based on objectively measurable IR-related traits and multimorbidity prevention strategies. These insights may pave the way for more targeted and effective treatments, ultimately enhancing patient outcomes.

## Ethical statement

We utilized publicly available summary statistics of genome-wide association studies conducted by external consortia and studies, and therefore no authorization was required from the local Ethics Committee.

## Data availability

All data are available upon request from the corresponding author.

## Acknowledgements

This publication is part of the project ’No labels needed: understanding psychiatric disorders based on genetic traits’ (with project number 09150161910091) of the research program Veni, which is partly financed by the Dutch Research Council (NWO) Health Research and Development (ZonMW). This project has received funding from the European Union’s Horizon 2020 research and innovation program under grant agreement No. 847879 (PRIME, Prevention and Remediation of Insulin Multimorbidity in Europe). The analyses were carried out on the Dutch national e-infrastructure with the support of SURF Cooperative. Research reported in this publication was supported by the National Institute Of Mental Health of the National Institutes of Health under Award Number R01MH124851.The content is solely the responsibility of the authors and does not necessarily represent the official views of any of the funders.

