## Supplementary Materials for "Unravelling the joint genetic architecture between psychiatic and insulin-related traits in the general population"

Linkage Disequilibrium Score Regression

To test genetic correlations for the inclusion criteria of our model we performed linkage disequilibrium score regression (LDSC) to obtain genetic correlations (rg) between the most recent ADHD GWAS (Demontis et al., 2023), ASD GWAS (Grove et al., 2019), and OCD GWAS (International Obsessive Compulsive Disorder Foundation Genetics & Studies, 2018) with the IR-related GWAS traits. The global genetic correlations were estimated as described on the publicly available GitHub page (https://github.com/bulik/ldsc) (Bulik-Sullivan et al., 2015). The LDSC analysis utilized linkage disequilibrium (LD) scores based on 1000 Genomes for GWASs of European descent (Genomes Project et al., 2015). The method involves two steps. First, the input summary statistics were munged to the appropriate LDSC format and filtered on info and MAF. Thereafter the genetic correlations are estimated. LDSC analysis to estimate genetic correlations was restricted to GWAS summary statistics with sample size N > 5 000. Given the exploratory phase of this study we included nominally significant genetic correlations in follow-up analyses. The observed genetic correlations between the aforementioned disorders and IR-related traits are summarized in Supplementary Table 1. We found nominally significant genetic correlations (p<0.05) for a total of five IR-related traits, namely BMI, FPG, FPI, HbA1c and HOMA-IR. In addition, compared to the study of Fanelli et al. (2022) we identified an additional nominally significant genetic correlation between ADHD and HOMA-IR (rg=0.1727, p=0.0218) based on the larger most recent ADHD GWAS.

**Table S1. Genetic correlations between ADHD, ASD, OCD with IR-related traits.** Observed genetic correlations values (rg (p-value)). Abbreviations: ADHD= attention-deficit/hyperactivity disorder; ASD= autism spectrum disorder; OCD= obsessive-compulsive disorder; 2hGlu= glucose levels two hours after an oral glucose challenge; BMI=body mass index; FPI=fasting insulin; FPG=fasting glucose; Hb1ac=glycated haemoglobin; HOMA-B=homeostatic model assessment of β-cell function; HOMA-IR=homeostatic model assessment for insulin resistance. *Nominally significant genetic correlation (p<0.05).

| Disorder/trait | 2hGlu | BMI | FPG | FPI | HbA1c | HOMA-B | HOMA-IR |
|---|---|---|---|---|---|---|---|
| ADHD | -0.0199 (0.6621) | 0.3312 (1.0956e-63)* | 0.1691 (7.0e-4)* | 0.1691 (1.0e-4)* | 0.1356 (2.0e-4)* | 0.0131(0.8447) | 0.1727 ( 0.0218)* |
| ASD | 0.0766 (0.2263) | 0.0432 (1.6397e-01) | -0.0433 (0.3329) | -0.0167(0.7972) | -0.0298 (0.6188) | 0.0209(0.8516) | 0.0260 (0.8172) |
| OCD | 0.1152 (0.1678) | -0.2839 (2.5725e-11)* | -0.0716 (0.3398) | -0.1076 (0.1981) | 0.0787 (0.3670) | -0.0807(0.4817) | -0.1387 (0.2552) |

**Table S2. Top 10 SNPs of GWAS summary statistics of ADHD-IR latent factor.** Top 10 significant SNPs of the input GWAS summary statistics of ADHD-IR latent factor after removal of significant Q SNPs. Genome wide significance threshold was set at p=5e−8. chr=chromosome; bp=base pair position; non_effect_allele=non-effect allele; effect_allele=effect allele; rsID=variant ID; p=p-value; beta=beta value; se=standard error.

| chr | bp | non_effect_allele | effect_allele | rsID | p | beta | se |
|---|---|---|---|---|---|---|---|
| 7 | 44216137 | A | G | rs2908290 | 1.23235416585e-07 | -0.0314242503397 | 0.00594185000273 |
| 7 | 44189274 | C | T | rs2268575 | 2.44219171632e-07 | -0.0370280035427 | 0.00717307639586 |
| 7 | 44247258 | T | C | rs3824065 | 2.93418582276e-07 | 0.0350439124426 | 0.00683433471187 |
| 7 | 44219074 | C | T | rs1303722 | 3.87266877872e-07 | -0.0256517553898 | 0.00505442107295 |
| 7 | 44245363 | G | A | rs10278336 | 3.99919946051e-07 | 0.0343050546169 | 0.00676762333728 |
| 7 | 44231216 | G | T | rs3757840 | 5.47842566292e-07 | 0.0343666015303 | 0.00686131097124 |
| 2 | 169766560 | T | C | rs17539351 | 6.18108202085e-07 | 0.0533601160781 | 0.0107031228088 |
| 2 | 169749841 | C | T | rs853770 | 7.04593355211e-07 | 0.03107472714 | 0.00626494812754 |
| 8 | 118185063 | G | C | rs2466294 | 7.51613270752e-07 | 0.0234829192695 | 0.00474639300434 |
| 7 | 44200884 | C | T | rs2041547 | 7.98747788905e-07 | -0.0232076666168 | 0.00470202663414 |

**Table S3. Gene-based top significant associations results for the ADHD-IR latent factor.** Genome wide significance threshold was set at p=0.05/16888=2.961e-6 accounting for the 16888 number of tested genes. CHR=chromosome; START=start position gene; STOP=end position of gene; NSNPS= number of SNPs; NPARAM=number of parameters; N=sample size; ZSTAT=Z-value for the gene based on its p-value; P=p-value.

| Ensembl ID | CHR | START | STOP | NSNPS | NPARAM | N | ZSTAT | P | GENE |
|---|---|---|---|---|---|---|---|---|---|
| ENSG00000196353 | 3 | 131252399 | 132004254 | 877 | 54 | 499492 | 6.8775 | 3.0464e-12 | CPNE4 |
| ENSG00000179915 | 2 | 50145643 | 51259674 | 1275 | 79 | 499492 | 6.3997 | 7.7826e-11 | NRXN1 |
| ENSG00000175806 | 8 | 9911778 | 10286401 | 562 | 37 | 499492 | 6.3043 | 1.4478e-10 | MSRA |
| ENSG00000175161 | 3 | 85008132 | 86123579 | 677 | 28 | 499492 | 6.1891 | 3.0255e-10 | CADM2 |
| ENSG00000134571 | 11 | 47352957 | 47374253 | 11 | 4 | 499492 | 5.9654 | 1.2199e-09 | MYBPC3 |
| ENSG00000141564 | 17 | 78518619 | 78940171 | 470 | 42 | 499492 | 5.6214 | 9.4726e-09 | RPTOR |
| ENSG00000185053 | 8 | 13947373 | 15095848 | 1484 | 98 | 499492 | 5.5857 | 1.164e-08 | SGCZ |
| ENSG00000110514 | 11 | 47290712 | 47351582 | 15 | 3 | 499492 | 5.5746 | 1.2404e-08 | MADD |

| | | | | | | | | | |
|---|---|---|---|---|---|---|---|---|---|
| ENSG00000198597 | 19 | 30719197 | 31204445 | 317 | 48 | 499492 | 5.5733 | 1.2501e-08 | ZNF536 |
| ENSG00000084734 | 2 | 27719709 | 27746554 | 7 | 2 | 499492 | 5.4315 | 2.7935e-08 | GCKR |
| ENSG00000078328 | 16 | 6069095 | 7763340 | 2640 | 260 | 499492 | 5.3952 | 3.4228e-08 | RBFOX1 |
| ENSG00000120341 | 1 | 177893091 | 177953438 | 74 | 14 | 499492 | 5.3754 | 3.8197e-08 | SEC16B |
| ENSG00000106633 | 7 | 44183872 | 44237769 | 26 | 7 | 499492 | 5.316 | 5.3049e-08 | GCK |
| ENSG00000153707 | 9 | 8314246 | 10612723 | 3112 | 282 | 499492 | 5.255 | 7.4016e-08 | PTPRD |
| ENSG00000205334 | 2 | 27928653 | 27938599 | 6 | 2 | 499492 | 5.2359 | 8.21e-08 | AC074091.13 |
| ENSG00000163618 | 3 | 62384022 | 62861054 | 503 | 56 | 499492 | 5.0931 | 1.7614e-07 | CADPS |
| ENSG00000148498 | 10 | 34398488 | 35104253 | 503 | 28 | 499492 | 5.0866 | 1.8228e-07 | PARD3 |
| ENSG00000145743 | 5 | 107194736 | 107717799 | 450 | 29 | 499492 | 5.0482 | 2.2296e-07 | FBXL17 |
| ENSG00000149187 | 11 | 47487496 | 47587121 | 21 | 5 | 499492 | 5.0463 | 2.2519e-07 | CELF1 |
| ENSG00000148943 | 11 | 27516123 | 27528320 | 3 | 1 | 499492 | 4.9976 | 2.9025e-07 | LIN7C |
| ENSG00000183117 | 8 | 2792875 | 4852494 | 4262 | 366 | 499492 | 4.9928 | 2.975e-07 | CSMD1 |
| ENSG00000185046 | 12 | 99120235 | 100378432 | 881 | 80 | 499492 | 4.9571 | 3.5782e-07 | ANKS1B |
| ENSG00000153820 | 2 | 228844666 | 229046361 | 170 | 18 | 499492 | 4.9094 | 4.5677e-07 | SPHKAP |
| ENSG00000164076 | 3 | 49895421 | 49907655 | 7 | 2 | 499492 | 4.9088 | 4.5809e-07 | CAMKV |
| ENSG00000140443 | 15 | 99192200 | 99507759 | 201 | 39 | 499492 | 4.8907 | 5.0252e-07 | IGF1R |
| ENSG00000183715 | 11 | 132284871 | 133402414 | 1094 | 146 | 499492 | 4.8646 | 5.7355e-07 | OPCML |
| ENSG00000122008 | 5 | 74807581 | 74896969 | 25 | 5 | 499492 | 4.8424 | 6.4139e-07 | POLK |
| ENSG00000154027 | 1 | 77747736 | 78025651 | 283 | 36 | 499492 | 4.8225 | 7.0894e-07 | AK5 |
| ENSG00000153495 | 13 | 111968531 | 111996596 | 29 | 8 | 499492 | 4.8155 | 7.343e-07 | TEX29 |
| ENSG00000135472 | 12 | 50260679 | 50298000 | 20 | 6 | 499492 | 4.8015 | 7.8731e-07 | FAIM2 |
| ENSG00000113163 | 5 | 74664311 | 74807963 | 29 | 5 | 499492 | 4.7483 | 1.0259e-06 | COL4A3BP |
| ENSG00000123444 | 11 | 47593749 | 47600567 | 1 | 1 | 499492 | 4.7392 | 1.0726e-06 | KBTBD4 |

| | | | | | | | | | |
|---|---|---|---|---|---|---|---|---|---|
| ENSG00000213619 | 11 | 47586888 | 47606114 | 1 | 1 | 499492 | 4.7392 | 1.0726e-06 | NDUFS3 |
| ENSG00000152253 | 2 | 169690642 | 169769881 | 71 | 8 | 499492 | 4.7355 | 1.0926e-06 | SPC25 |
| ENSG00000213996 | 19 | 19375173 | 19384200 | 2 | 1 | 499492 | 4.7313 | 1.1155e-06 | TM6SF2 |
| ENSG00000150672 | 11 | 83166055 | 85338966 | 1631 | 113 | 499492 | 4.721 | 1.1736e-06 | DLG2 |
| ENSG00000133958 | 14 | 93799565 | 94174222 | 170 | 12 | 499492 | 4.7195 | 1.182e-06 | UNC79 |
| ENSG00000155093 | 7 | 157331750 | 158380480 | 526 | 90 | 499492 | 4.7163 | 1.2009e-06 | PTPRN2 |
| ENSG00000185352 | 13 | 96743093 | 97485671 | 439 | 33 | 499492 | 4.7148 | 1.2096e-06 | HS6ST3 |
| ENSG00000164078 | 3 | 49924435 | 49941299 | 6 | 2 | 499492 | 4.7118 | 1.2276e-06 | MST1R |
| ENSG00000165916 | 11 | 47440320 | 47447993 | 6 | 2 | 499492 | 4.7039 | 1.2759e-06 | PSMC3 |
| ENSG00000134574 | 11 | 47236493 | 47260767 | 8 | 4 | 499492 | 4.6616 | 1.5687e-06 | DDB2 |
| ENSG00000066032 | 2 | 79412357 | 80875905 | 1678 | 169 | 499492 | 4.6572 | 1.603e-06 | CTNNA2 |
| ENSG00000091844 | 6 | 153325594 | 153452384 | 136 | 12 | 499492 | 4.6564 | 1.6086e-06 | RGS17 |
| ENSG00000109920 | 11 | 47738072 | 47788995 | 14 | 2 | 499492 | 4.6535 | 1.632e-06 | FNBP4 |
| ENSG00000069667 | 15 | 60780483 | 61521518 | 733 | 118 | 499492 | 4.6404 | 1.7385e-06 | RORA |
| ENSG00000184349 | 5 | 106712590 | 107006596 | 269 | 43 | 499492 | 4.6328 | 1.8042e-06 | EFNA5 |
| ENSG00000083067 | 9 | 73143979 | 74061820 | 863 | 67 | 499492 | 4.6165 | 1.9515e-06 | TRPM3 |
| ENSG00000166436 | 11 | 8633584 | 8693413 | 33 | 3 | 499492 | 4.6154 | 1.9614e-06 | TRIM66 |
| ENSG00000165923 | 11 | 47681143 | 47736941 | 16 | 3 | 499492 | 4.6031 | 2.081e-06 | AGBL2 |
| ENSG00000186094 | 1 | 48998527 | 50489585 | 619 | 31 | 499492 | 4.5986 | 2.127e-06 | AGBL4 |
| ENSG00000136944 | 9 | 129376722 | 129463311 | 69 | 15 | 499492 | 4.588 | 2.2372e-06 | LMX1B |
| ENSG00000030066 | 11 | 47799639 | 47870107 | 25 | 3 | 499492 | 4.5866 | 2.2531e-06 | NUP160 |
| ENSG00000187391 | 7 | 77646393 | 79082890 | 1560 | 159 | 499492 | 4.538 | 2.84e-06 | MAGI2 |
| ENSG00000066382 | 11 | 30406040 | 30608419 | 212 | 22 | 499492 | 4.5365 | 2.8596e-06 | MPPED2 |
| ENSG00000145934 | 5 | 166711804 | 167691162 | 727 | 97 | 499492 | 4.5356 | 2.8717e-06 | TENM2 |

| | | | | | | | | | |
|---|---|---|---|---|---|---|---|---|---|
| ENSG00000104447 | 8 | 116420724 | 116821899 | 234 | 12 | 499492 | 4.5309 | 2.9367e-06 | TRPS1 |

**Table S4. Top 10 SNPs of GWAS summary statistics of OCD/OCS-IR latent factor.** Top 10 significant SNPs of the input GWAS summary statistics of the OCD/OCS-IR latent factor after removal of significant Q SNPs. Genome wide significance threshold was set at p=5e−8. chr=chromosome; bp=base pair position; non_effect_allele=non-effect allele ; effect_allele=effect allele ; rsID=variant ID; p=p-value; beta=beta value; se=standard error.

| chr | bp | non_effect_allele | effect_allele | rsID | p | beta | se |
|---|---|---|---|---|---|---|---|
| 11 | 92725321 | G | A | rs6483213 | 4.26666647837e-07 | 0.0605271100128 | 0.0119697843957 |
| 11 | 92721787 | C | A | rs4406791 | 4.45707923015e-07 | 0.0605789770861 | 0.0119998247759 |
| 11 | 92715849 | A | G | rs1562444 | 4.77044447141e-07 | 0.0619143591377 | 0.012296006151 |
| 11 | 92718163 | G | A | rs1447351 | 4.80849109092e-07 | 0.0604918284528 | 0.0120171277617 |
| 11 | 92721319 | A | G | rs1597023 | 4.99532003042e-07 | 0.0606171892831 | 0.0120595403749 |
| 11 | 92719207 | G | T | rs4611171 | 5.11057876668e-07 | 0.0591955688757 | 0.0117869819086 |
| 11 | 92722761 | A | G | rs1447352 | 5.40120894988e-07 | 0.0605033837903 | 0.012072949452 |
| 11 | 92717405 | A | C | rs4753072 | 5.46647872215e-07 | 0.0604188644844 | 0.0120616477213 |
| 11 | 92718127 | C | G | rs1447350 | 5.80757824249e-07 | 0.060014640605 | 0.0120089121843 |
| 11 | 92717475 | A | G | rs4753073 | 6.41973308734e-07 | 0.0601646989425 | 0.0120857723396 |

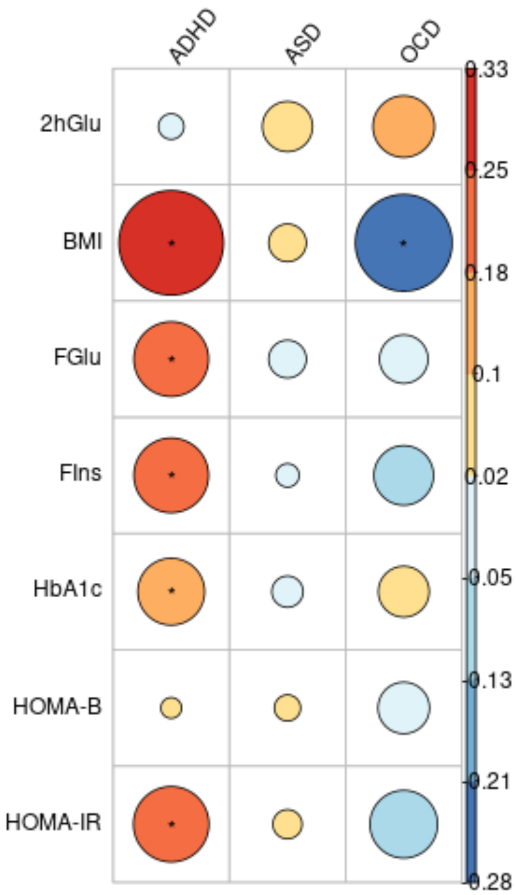


**Figure S1. Genetic correlation plot.** The circle represents the level of genetic correlations and the colors show the direction of correlation (warmer colors indicating a positive direction and cooler colors indicating a negative direction). Abbreviations: ADHD= attention-deficit/hyperactivity disorder; ASD= autism spectrum disorder; OCD= obsessive-compulsive disorder; 2hGlu= glucose levels two hours after an oral glucose challenge; BMI=body mass index; FPI=fasting insulin; FPG=fasting glucose; HbA1c=glycated haemoglobin; HOMA-B=homeostatic model assessment of β-cell function; HOMA-IR=homeostatic model assessment for insulin resistance. *Nominally significant genetic correlation ($p<0.05$).

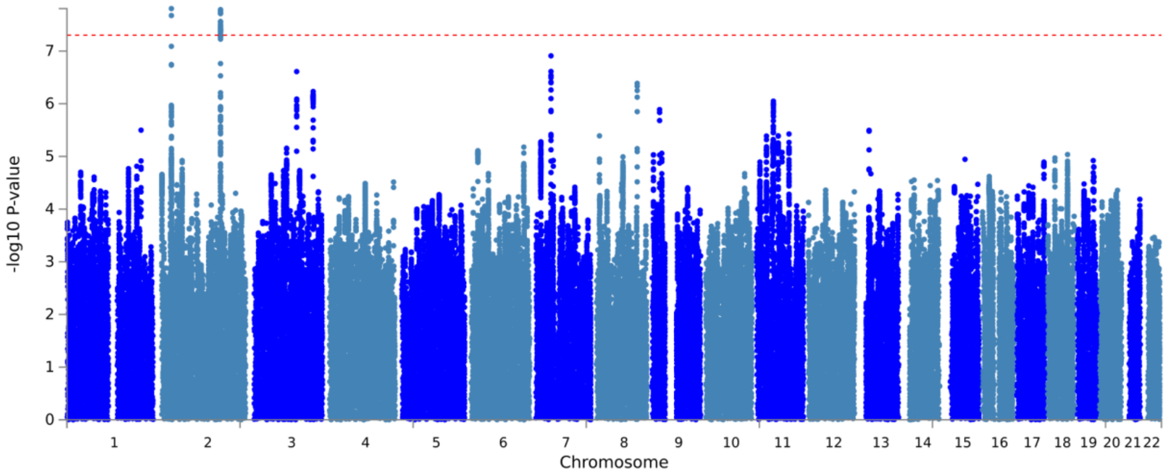


**Figure S2. Manhattan plot of GWAS summary statistics of ADHD-IR latent factor before removal of significant Q SNPs.** The x-axis displays the chromosomes, and the y-axis displays the −log p-value. The red dashed line in the plot is the genome-wide significance threshold (p=5e-8).

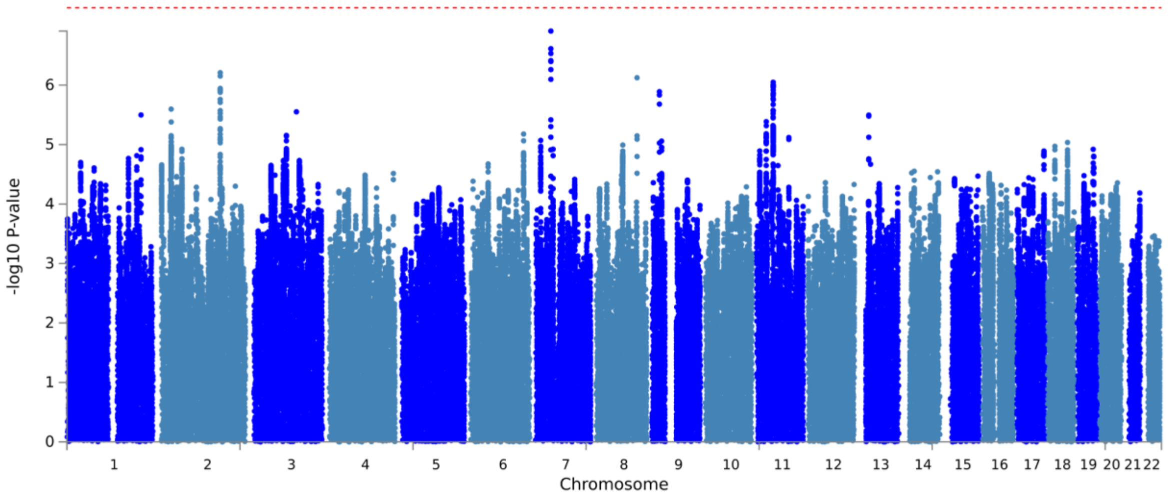


**Figure S3. Manhattan plot of GWAS summary statistics of ADHD-IR latent factor after removal of significant Q SNPs.** The x-axis displays the chromosomes, and the y-axis displays the −log p-value. The red dashed line in the plot is the genome-wide significance threshold (p=5e-8).

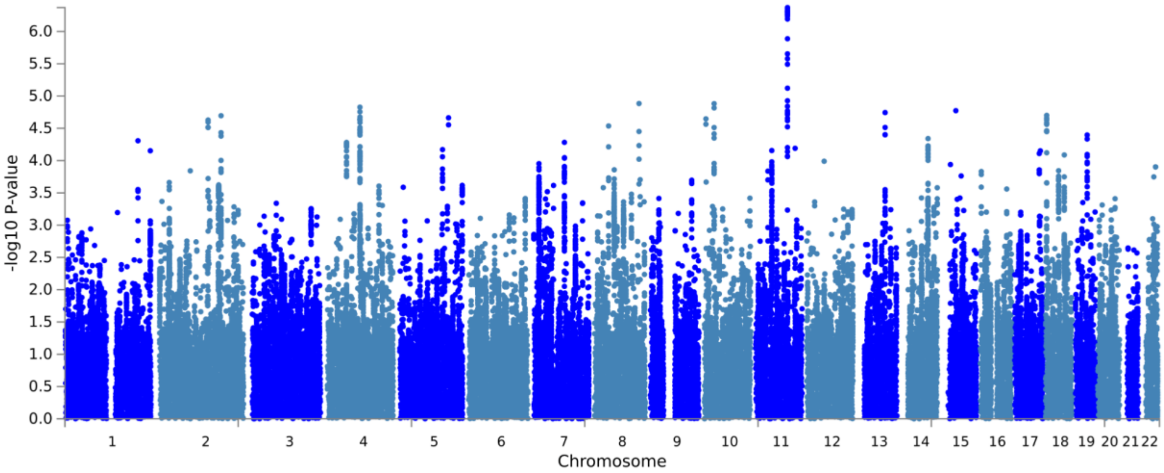


**Figure S4. Manhattan plot of GWAS summary statistics of OCD/OCS-IR latent factor before removal of significant Q SNPs.** The x-axis displays the chromosomes, and the y-axis displays the −log p-value. The genome-wide significance threshold (p=5e-8).

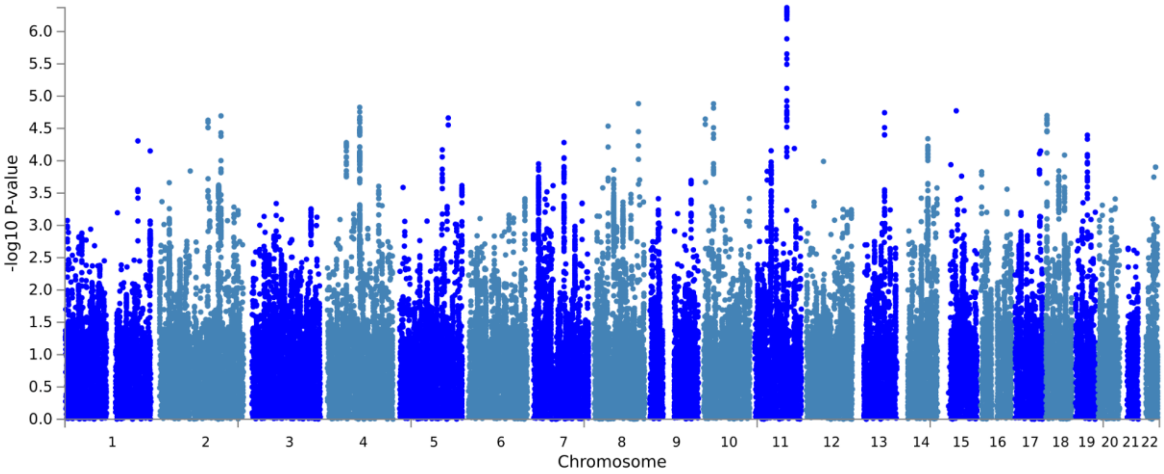


**Figure S5. Manhattan plot of GWAS summary statistics of OCD/OCS-IR latent factor after removal of significant Q SNPs.** The x-axis displays the chromosomes, and the y-axis displays the −log p-value. The genome-wide significance threshold (p=5e-8).

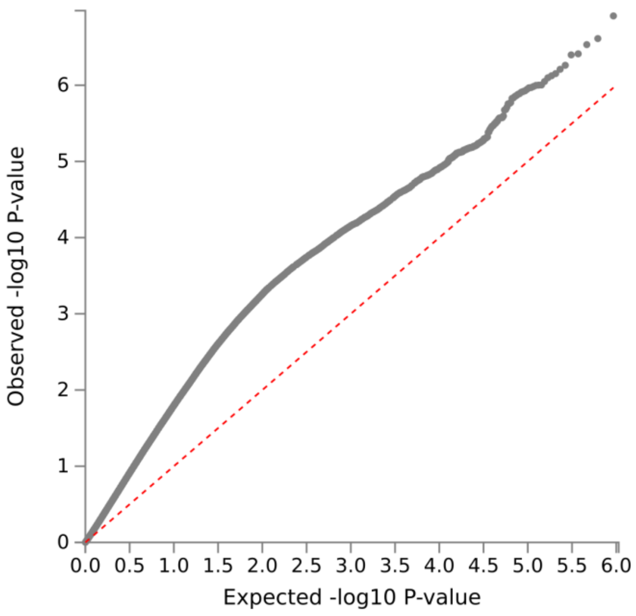


A

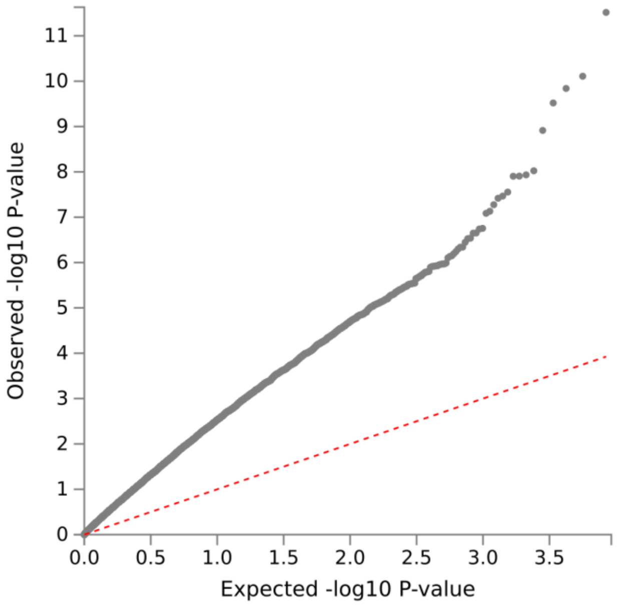


B

**Figure S6. Q-Q plots of ADHD-IR latent factor.** Quantile-quantile (Q-Q) plot of GWAS summary statistics of ADHD-IR latent factor after removal of significant Q SNPs (top; A) Q-Q plot of gene-based test of ADHD-IR latent factor after removal of significant Q SNPs. (bottom; B) The x-axis displays the expected −log p-value, and the y-axis displays the observed p-value.

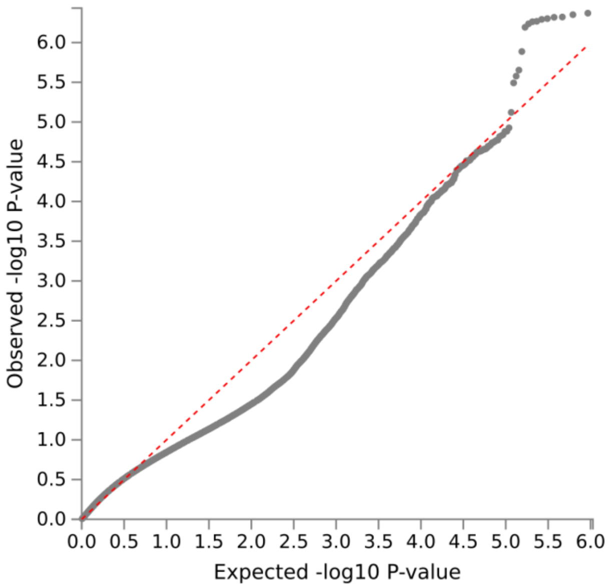


A

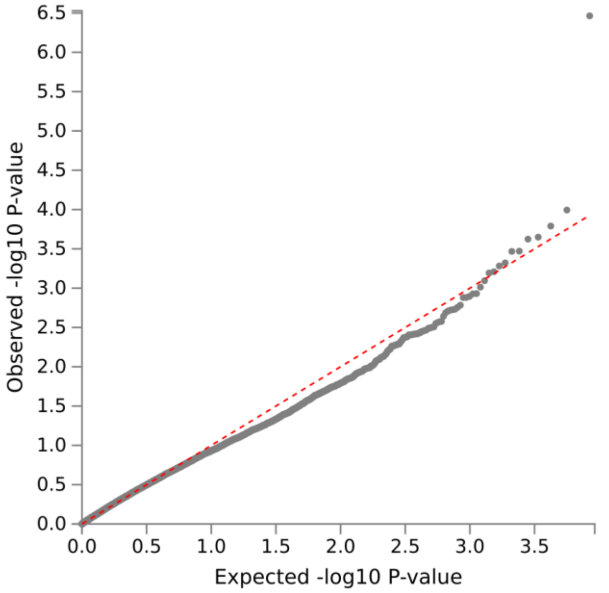


B

**Figure S7. Q-Q plots of OCD/OCS-IR latent factor.** Quantile-quantile (Q-Q) plot of GWAS summary statistics of OCD/OCS-IR latent factor after removal of significant Q SNPs (top;A). Q-Q plot of gene-based test of OCD/OCS-IR latent factor after removal of significant Q SNPs (bottom;B). The x-axis displays the expected −log p-value, and the y-axis displays the observed p-value.

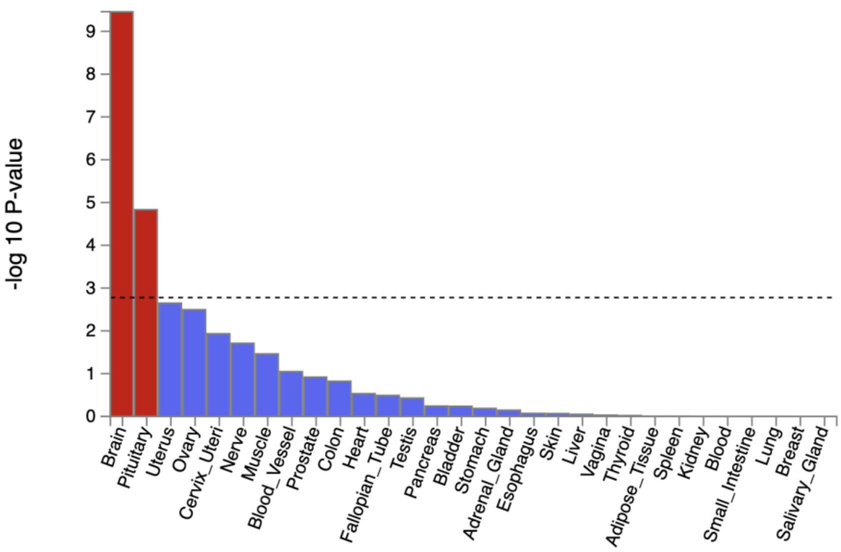


A

.

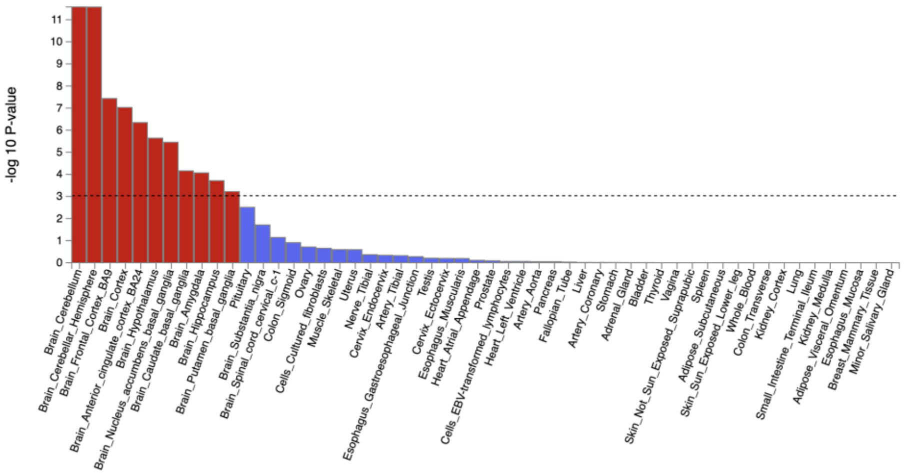


B

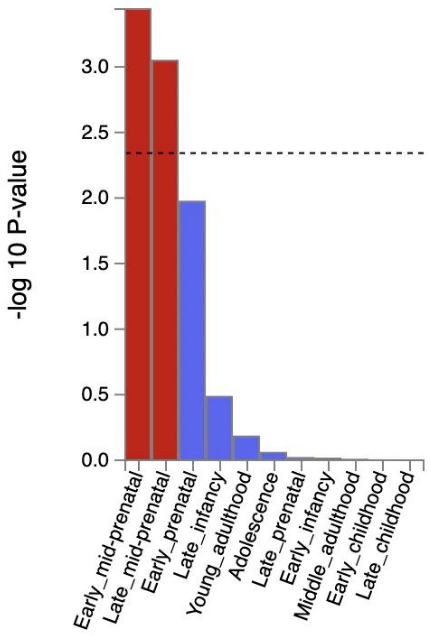


C

**Figure S8. MAGMA tissue expression analyses of tissue specificity of the genes related to ADHD-IR latent factor.** MAGMA gene-property analysis results for gene expression of the selected GTEx/v8 30 general tissue types (top;A), GTEx/v8 53 specific tissue types (middle;B) and BrainSpan (bottom;C) gene expression datasets.

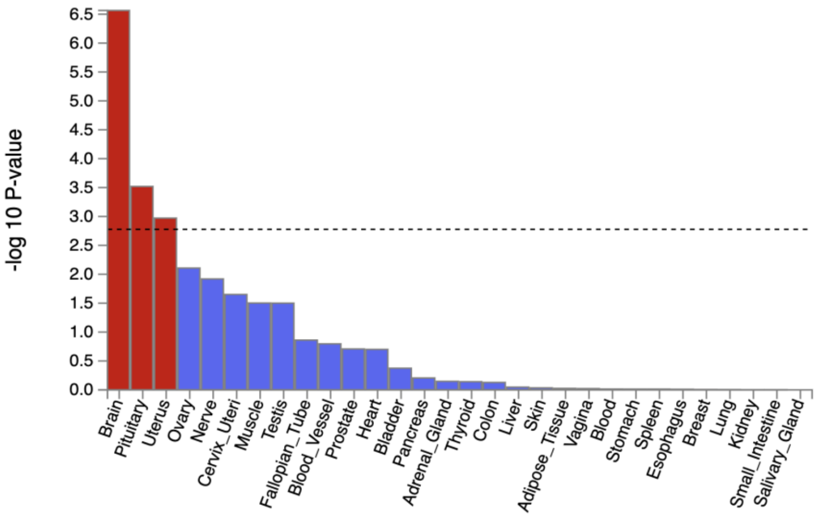
-log 10 P-value
6.5
6.0
5.5
5.0
4.5
4.0
3.5
3.0
2.5
2.0
1.5
1.0
0.5
0.0
Brain
Pituitary
Uterus
Ovary
Nerve
Cervix_Uteri
Muscle
Testis
Fallopian_Tube
Blood_Vessel
Prostate
Heart
Bladder
Pancreas
Adrenal_Gland
Thyroid
Colon
Liver
Skin
Adipose_Tissue
Vagina
Blood
Stomach
Spleen
Esophagus
Breast
Lung
Kidney
Small_Intestine
Salivary_Gland

A

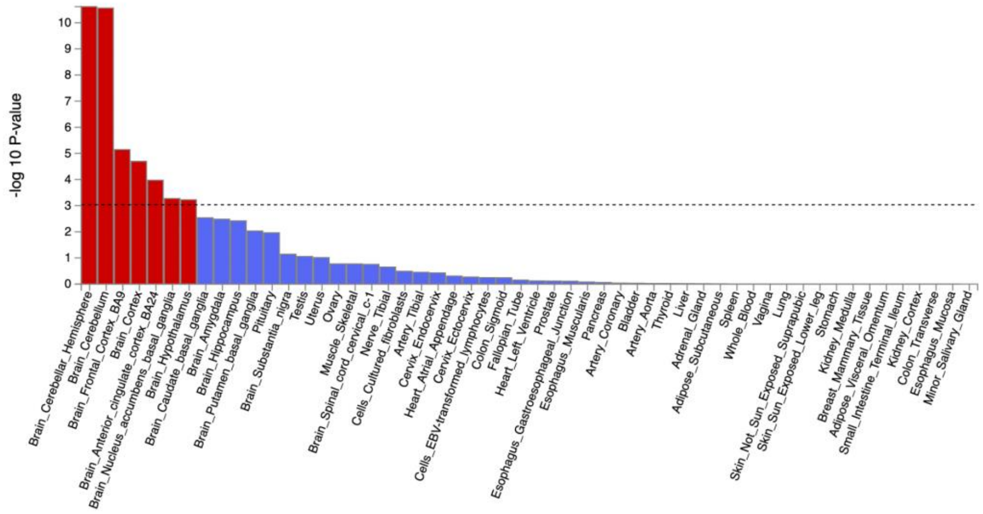
-log 10 P-value
10
9
8
7
6
5
4
3
2
1
0
Brain_Cerebellar_Hemisphere
Brain_Cerebellum
Brain_Frontal_Cortex_BA9
Brain_Cortex
Brain_Anterior_cingulate_cortex_BA24
Brain_Nucleus_accumbens_basal_ganglia
Brain_Hypothalamus
Brain_Caudate_basal_ganglia
Brain_Amygdala
Brain_Hippocampus
Brain_Putamen_basal_ganglia
Pituitary
Brain_Substantia_nigra
Testis
Uterus
Ovary
Muscle_Skeletal
Brain_Spinal_cord_cervical_c-1
Nerve_Tibial
Cells_Cultured_fibroblasts
Artery_Tibial
Cervix_Endocervix
Heart_Atrial_Appendage
Cervix_Ectocervix
Cells_EBV-transformed_lymphocytes
Colon_Sigmoid
Fallopian_Tube
Heart_Left_Ventricle
Prostate
Esophagus_Gastroesophageal_Junction
Esophagus_Muscularis
Pancreas
Artery_Coronary
Bladder
Artery_Aorta
Thyroid
Liver
Adrenal_Gland
Adipose_Subcutaneous
Spleen
Whole_Blood
Vagina
Lung
Skin_Not_Sun_Exposed_Suprapubic
Skin_Sun_Exposed_Lower_leg
Stomach
Kidney_Medulla
Breast_Mammary_Tissue
Adipose_Visceral_Omentum
Small_Intestine_Terminal_Ileum
Kidney_Cortex
Colon_Transverse
Esophagus_Mucosa
Minor_Salivary_Gland

B

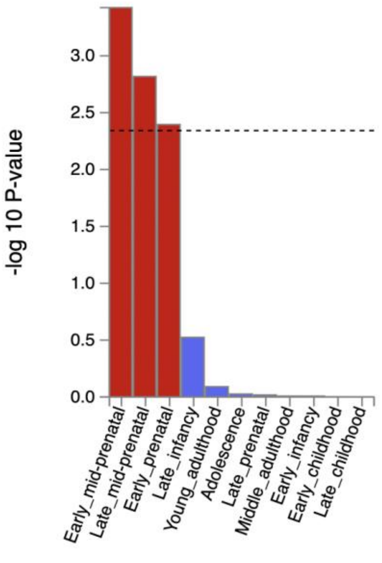
-log 10 P-value
3.0
2.5
2.0
1.5
1.0
0.5
0.0
Early_mid-prenatal
Late_mid-prenatal
Early_prenatal
Late_infancy
Young_adulthood
Adolescence
Late_prenatal
Middle_adulthood
Early_infancy
Early_childhood
Late_childhood

C

**Figure S9. MAGMA tissue expression analyses of tissue specificity of the genes related OCD/OCS-IR latent factor.** MAGMA gene-property analysis results for gene expression of the selected GTEx/v8 30 general tissue types (top;A), GTEx/v8 53 specific tissue types (middle;B) and BrainSpan (bottom;C) gene expression datasets.
